# Tricuspid Valve Annulus Size by Echocardiography: Predictor of Cardiac Limitation in Pediatric Pectus Excavatum

**DOI:** 10.1101/2024.02.12.24302736

**Authors:** James Chang, James Eubanks, Tim Jancelewicz, Vijaya Joshi, Hugo Martinez, Samir Shah, Elizabeth Paton, Ranjit Philip

## Abstract

**Background:** The severity of pectus excavatum (PEX) as measured by Haller index (HI) does not always correlate with symptoms of aerobic capacity. Transthoracic echocardiograms (TTE) are generally reported as normal which may influence the pediatrician’s decision to refer for corrective surgery. The aim of this study was to find a reproducible TTE marker as an indicator of right ventricular compression and compare it to severity of PEX and cardiopulmonary exercise test (CPET) indices.

**Methods:** The study included patients aged 10-19 years with an institution-based protocol for preoperative PEX evaluation with TTE, chest computed tomography (CT) for HI, and CPET from 2015-2021. We divided the patients into two groups, mild/moderate PEX (HI 2-3.5) and severe PEX (HI > 3.5). Tricuspid valve annulus size (TVAS) was compared between the groups as well as with other CPET and TTE indices using Student’s t-test. Spearman’s rank correlation coefficient was used to evaluate correlations between the severity of PEX by HI with the TTE and CPET parameters.

**Results:** Of the 124 patients, 82 (66.1%) had severe PEX and 42 (33.9%) had mild/moderate PEX. The mean TVAS z-scores in the mild/moderate PEX group was -1.98(SD 0.51) and -2.24 (SD 0.71) in the severe PEX group (p 0.046). There was a negative correlation between the TVAS z-score and the severity of PEX but this was not statistically significant (r = -0.154, p = 0.087). There was no significant difference in peak oxygen uptake (peak V_O2_) or left ventricular ejection fraction between the severity groups. However, the TVAS z-score positively correlated with peak V_O2_ (median 43 ml/kg/min, r = 0.023, p = 0.01), peak V_O2_ percent predicted (median 86%, r = 0.19, p = 0.04), and O_2_ pulse (median 12.7 ml/beat, r = 0.20, p = 0.025), and negatively correlated with V_E_/V_CO2_ (median 29, r = -0.23, p = 0.01).

**Conclusion:** The severity of PEX by HI does not factor in the location of cardiac compression and may not always reflect the degree of cardiac limitation. The Tricuspid valve annulus size is a good TTE indicator of cardiopulmonary compromise from PEX. A TVAS z score <-2 is a good predictor of cardiac compromise in pediatric PEX. This may provide additional functional parameters in the decision-making process for corrective surgery.

## Background

Pectus excavatum (PEX), or “funnel chest”, is a congenital disease where the anterior wall of the thoracic cavity is deformed posteriorly to varying degrees of severity (1). It is the most common type of pediatric chest wall deformity, accounting for over 90% of lesions. It occurs in 1 in 400 live births and disproportionately affects males more than females (2). The severity of disease is defined by the radiographic Haller Index (HI), which is a ratio of the transverse diameter of the thoracic cavity by the smallest distance between the anterior aspect of the vertebral body and the posterior aspect of the sternum on axial imaging (Figure. 1). A normal HI is less than 2 and severe is a HI greater than 3.5 (3). Widely-used objective criteria for surgical PEX repair are a HI >3.25, cardiac compression or displacement, mitral valve prolapse, conduction abnormalities, pulmonary function testing showing restrictive respiratory disease, or a failed previous repair (4-6).

**Figure 1.**
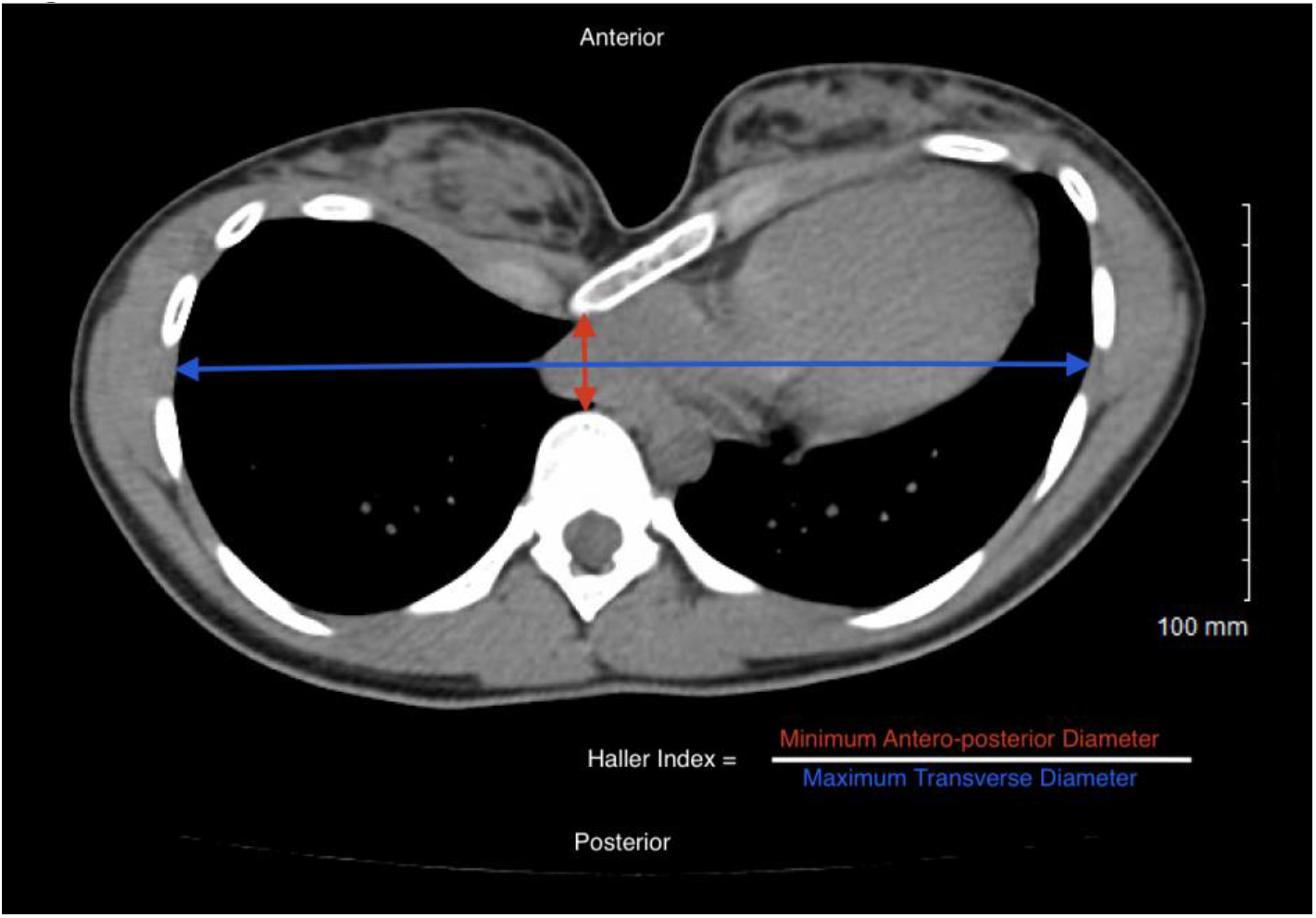
Axial plane of a CT scan in a patient with pectus excavatum demonstrating the Haller Index. There is clear compression of the right-sided cardiac structures and mass effect on the heart, shifting the cardiac mass into the left-side of the chest.

Generally, patients with PEX report cardiopulmonary symptoms with exertion and exercise impairment but they can be asymptomatic as well. Reported symptoms include chest pain, tachycardia, palpitations, exercise intolerance, dyspnea on exertion, exercise-induced wheezing, syncope, dizziness, light-headedness, and significant psycho-social symptoms such as anxiety and body image disturbance (7). Frequently, these symptoms do not correlate with the severity of PEX as measured by HI (8). It is theorized that the posterior displacement and deformation of the sternal wall can cause compression of the right-sided cardiac structures, which leads to functional impairment (3, 4, 7-11). Unfortunately, the HI as a metric of severity does not take into account the location of the PEX or the degree of compression of cardiopulmonary structures but moreso the severity of the skeletal deformity.

Transthoracic echocardiography (TTE) and cardiopulmonary exercise testing (CPET) are standard modalities used during pre-operative evaluation of PEX. Based on anecdotal experience as well as in literature reviews and meta-analysis, it is notable that the baseline measured resting echocardiographic variables for function are largely within normal limits. This is likely secondary to the slow progressive nature of the disease and cardiac adaptation at rest (9, 12).

This has made pediatricians question the need for PEX repair in children with “normal” clinical testing. There have been studies that investigate markers of right ventricular changes in PEX, but it is difficult to assess the right-sided structures on TTE due to the deformation of the anterior chest wall as well the complex geometry of the right ventricle (13, 14). Despite these limitations, TTE is integral in the cardiac assessment in PEX patients.

Our aim was to find a pragmatic, reliable and reproducible echocardiographic marker for the general cardiologist that may aid the clinician in decision-making for PEX repair. The goal was to use a TTE marker that would be a reproducible measure for right ventricular compression from the PEX. To test this, the TTE marker would have to be compared with the severity of PEX by HI and metabolic markers on a CPET. We hypothesize that the tricuspid valve annulus size (TVAS) as a measure of right ventricular compression, is negatively correlated with the severity of PEX by HI. The primary outcome of the study was the correlation between TVAS and HI. The secondary outcome was comparison of TVAS to metabolic markers on CPET.

## Methodology

### Study Design

A single center, retrospective review was performed on all patients diagnosed with PEX who underwent an institutional protocoled preoperative evaluation at the Le Bonheur Children’s Heart Institute in Memphis, Tennessee between January 2015 and December 2021. The protocol included clinical assessment with physical exam, TTE, radiography [computed tomography (CT) of the thorax or chest x-ray], and a CPET utilizing the Bruce Protocol as recommended by the guidelines for exercise testing in the pediatric population per the American Heart Association (15). The study involved the review of the electronic medical record as well as re-evaluation of archived echocardiographic images. The University of Tennessee Institutional Review Board waived the need for parental consent and approved this study.

### Study Population

Patients who were referred to the Heart Institute for a protocoled evaluation for PEX were included in the study. Patients were excluded if there was another cause for chest wall deformity (pectus carinatum, trauma, and thoracic procedures or if they had congenital structural intra-cardiac abnormalities.

#### Definitions

##### 1. Haller Index

The severity of PEX was stratified utilizing the HI measured by radiography as part of the preoperative evaluation. The HI was stratified into mild/moderate (2-3.5) and severe (>3.5) categories (Figure 1).

##### 2. Echocardiographic Parameters

The Le Bonheur Children’s Heart Institute echocardiography lab is accredited by the Inter-societal Accreditation Commission (IAC) and routine echocardiograms are performed in keeping with the guidelines and standards for performance of pediatric echocardiograms set forth by the American Society of Echocardiography. Echocardiogram images were independently reviewed by a pediatric cardiologist.

###### Tricuspid Valve Annulus Size (TVAS)

The TVAS (mm) was measured at end-diastole in the modified apical four-chamber view to maximize right heart size. The standards for the modified apical four-chamber view were: 1.) clear visualization of the tricuspid valve left of the center of frame; 2.) best visualization of the right ventricular free wall, apex, and septum; and 3.) visualization of the right atrial and ventricular chambers. End-diastole was defined as the frame at which the atrioventricular valve has closed with corresponding electrocardiogram tracing at the peak of the QRS wave (see Figure 2) (16). The TVAS z-score was calculated based on the individual’s body surface area and compared to the Detroit Data set (17). To test reproducibility and inter-observer variability in measurements, 3 pediatric cardiologists (senior, mid-career and a pediatric cardiology fellow in training) reviewed these images and made independent TVAS measurements based on the criteria above.

**Figure 2:**
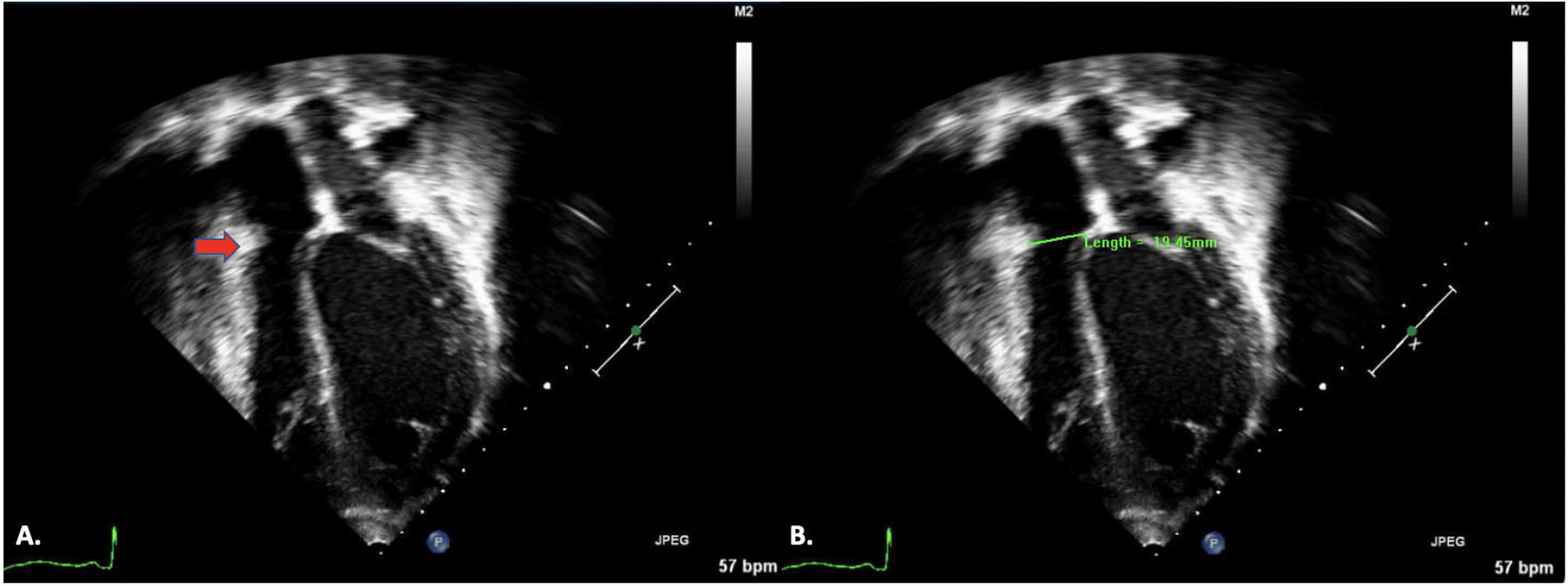
Modified Apical 4-Chamber View by transthoracic echocardiogram in a patient with pectus excavatum. A.) The right ventricular free wall and tricuspid valve annulus (red arrow) are significantly compressed in this view. B.) The TVAS as measured in this modified apical four-chamber view at end-diastole.

###### Other Echocardiographic Parameters

In addition to TVAS, other measurable data taken from the TTE study were cardiac output by left ventricular outflow tract velocity time integral (LVOT VTI in L/min), fractional area of change of the right ventricle (FAC in %) obtained in the standard right-ventricle-focused four-chamber apical view, the ejection fraction by motion-mode echocardiography (LVEF in %), and the following tissue Doppler measurements: tricuspid annular plane systolic excursion (TAPSE in mm); left ventricle E’ lateral [in centimeters per second (cm/s)]; E’ septal (cm/s); and the lateral right ventricular wall defined as E’ RV (cm/s).

##### 3. Cardiopulmonary Parameters

We reviewed the CPET data for several metabolic markers that are widely used for assessment of cardiopulmonary and exercise capacity. This includes oxygen consumption (peak V_O2_), percent-predicted maximum oxygen consumption (peak V_O2_ percent-predicted), oxygen pulse at anaerobic threshold (O_2_ pulse), percent-predicted oxygen pulse (O_2_ pulse percent-predicted), ventilatory efficiency (V_E_/V_CO2_), resting heart rate (beats per minute (bpm)), heart rate reserve (HRR in bpm), and 2-minute heart rate recovery (bpm).

a. The *peak V*_*O2*_ is measured in milliliters per kilogram per minute (ml/kg/min), and is defined as the maximum amount of oxygen uptake by skeletal muscles during exercise, measured in volume of oxygen per heart beat (ml O_2_/beat) and correlates with maximum aerobic capacity. A lower peak V_O2_ can be associated with functional cardiac disease and inability to meet the metabolic demands of the body during exercise (18). Percent-predicted peak V_O2_ is the percent of the expected peak V_O2_ achieved for age, weight, and gender by the patient and is used in similar fashion as the measured peak V_O2_.
b. The *O*_*2*_ *pulse* is the ratio of oxygen consumption to the heart rate and is a surrogate for the ability of the heart to deliver oxygen/cardiac output during maximal aerobic exercise. The O_2_ pulse percent-predicted is the percent of the expected O_2_ pulse achieved for age, weight, and gender.
c. The *V*_*E*_*/V*_*CO2*_ is the ratio the minute ventilation over carbon dioxide output and reflects ventilator efficiency.
d. *Heart rate reserve* in CPET testing is the difference between the maximum predicted HR and the observed maximum HR. A normal value is < 15bpm (19). A high HRR may indicate a sub-maximal effort.
e. The *2-minute heart rate recovery* is the difference between the observed maximum heart rate and the heart rate after 2 minutes of rest immediately following maximal effort. It is a measure of fitness and conditioning i.e. a higher heart rate recovery indicates better conditioning and fitness. The average 1 and 2-minute heart rate recovery in elite adolescent athletes is 23 bpm and 58 bpm respectively (20).

###### Data Analysis

We divided the patients into two groups, mild/moderate PEX (HI 2 to 3.5) and severe PEX (HI greater than 3.5). The 2 groups were compared to evaluate for potential significant differences by disease severity, defined by the HI. The mean and standard deviations of TTE and CPET markers for the 2 groups were calculated and the groups were compared using independent samples t-test. Mann-Whitney U tests for group comparisons of variables that violated statistical assumptions was used. Spearman’s rank correlation coefficient was used to evaluate correlations between the severity of PEX by HI with the TTE and CPET parameters. In addition, the TVAS z-score was also independently evaluated for correlation with TTE and CPET parameters. A p-value of ≤ 0.05 was considered statistically significant. Data analysis was performed utilizing the SPSS software Version 25 for all statistical analysis (SPSSS, Chicago, IL).

## Results

There were 124 patients in the cohort, which was predominantly male (105, 84.7%). The median age was 15 years (range: 7-19). Of the 124 patients, 42 had mild/moderate PEX (median 3.2, IQR 2.9-3.4) and 82 had severe PEX (median 4.5, IQR 3.7-5.35) as shown in Table 1. There were 27 asymptomatic PEX patients (22%). Among the 78% that were symptomatic, the most common symptoms reported were respiratory symptoms of dyspnea on exertion or shortness of breath and chest pain (Table 2).

**Table 1:** Demographic Data of the Study Population with grouped severity based on Haller Index.

| Demographic Data |  |
| --- | --- |
| Age (years), range (median) | 7-19 (15) |
| Male, n (%) | 105 (84.7%) |
| Female, n (%) | 19 (15.3%) |
| Height (cm), range (median) | 122.5 – 195 (173) |
| Weight (kg), range (median) | 23.4 – 90 (56) |
| BMI (m<sup>2</sup>), range (median) | 13.8 – 27.5 (18.7) |
| BSA (kg/m<sup>2</sup>), range (median) | 0.89 – 2.2 (1.6) |
| Haller index (HI) , range (median) | 2.35 – 20 (3.7) |
| Mild & moderate PEX: HI n (median HI, IQR) | 42 (3.2, 2.9-3.4) |
| Severe PEX: HI n (median HI, IQR) | 82 (4.5, 3.7-5.35) |

**Table 2:** The distribution of symptoms among the symptomatic patients in the study group.

| Symptoms | Symptom Distribution (% , n) |
| --- | --- |
| Respiratory Symptoms | 60% (n = 58) |
| Chest Pain | 54% (n = 52) |
| Exercise Intolerance | 39% (n = 38) |
| Syncope/Pre-syncope | 24% (n = 23) |
| Palpitations/Tachycardia | 23% (n = 22) |

### Echocardiographic Parameters

The TVAS z-score was lower across both groups of PEX in comparison to normal values for age: median z-score for mild/moderate PEX was -2 (IQR: -1.5 to -2.4) and was -2.2 (IQR : -1.6 to -2.8) for severe PEX (Table 3). There was no significant difference in the absolute TVAS (p=0.25) but the mean TVAS z-scores between the 2 groups of severity were statistically significant (p= 0.046). As expected, there was a negative correlation between the TVAS z-score and the severity of PEX but this was not statistically significant (r = -0.154, p = 0.087) as shown in Table 4. There was no significant correlation between TVAS and the other TTE markers. There was a very good strength of agreement between the cardiologists in measurement of the TVAS (Fleiss’ kappa value of 0.85).

**Table 3:** Comparison of the echocardiographic measurements by transthoracic echocardiography and the metabolic markers on cardiopulmonary testing between the two severity groups of pectus excavatum.

| Echocardiographic<br>&<br>Cardiopulmonary<br>Exercise Test<br>Parameters | Mild & Moderate PEX<br>(HI= 2-3.5) |  | Severe PEX<br>(HI>3.5) |  | P-value |
| --- | --- | --- | --- | --- | --- |
|  | *Median IQR* | *Mean (SD)* | *Median IQR* | *Mean (SD)* |  |
| TVAS (cm) | 2.11(1.94 - 2.26) | 2.1(0.31) | 2.01 (1.79 - 2.25) | 2.03(0.34) | 0.251 |
| TVAS z-score | -2.04 (-2.41 - -1.54) | -1.98(0.51) | -2.23 (-2.77 - -1.57) | -2.24 (0.71) | 0.046* |
| LVEF (%) | 64 (61 - 68) | 64(5) | 63 (58 - 66) | 62(6) | 0.167 |
| Cardiac Index<br>(L/min/m<sup>2</sup>) | 2.44 (1.82 - 2.93) | 2.33 (0.60) | 2.19 (1.84-2.53) | 2.24 (0.58) | 0.415 |
| E' lateral (cm/s) | 16.88 (13.5 - 18.63) | 16.55 (4.06) | 16.6 (11.7 - 19.85) | 16.35 (5.09) | 0.829 |
| E' septal (cm/s) | 12.95 (11.2 - 14.53) | 13.12(3.31) | 12.5 (10.65 - 14.75) | 12.65(2.99) | 0.443 |
| E' RV (cm/s) | 14.4 (11.7 - 16.3) | 13.58(3.28) | 14.49 (11.67 - 16.81) | 14.01 (3.53) | 0.578 |
| TAPSE (mm) | 22.3 (18.3-23.8) | 21.31(3.73) | 20.3(17.6-23.1) | 20.89 (3.87) | 0.602 |
| V<sub>E</sub>/V<sub>CO2</sub> | 29 (27 - 31) | 30.1 (4.19) | 30 (28 - 31) | 29.85 (7.04) | 0.694 |
| **Peak V<sub>O2</sub> (ml/kg/min)** | 43.4 (38.35 – 47.78) | 41.9 (7.59) | 43.05 (37.35 – 48.55) | 43.13(8.97 | 0.464 |
| **Peak V<sub>O2</sub> Percent Predicted (%)** | 85.5 (74 – 93.5) | 82.96 (16.58) | 85.5 (72 – 98) | 86.4 (18.52) | 0.327 |
| **O<sub>2</sub> Pulse (ml O<sub>2</sub> /beat)** | 13.4 (10.65 – 15) | 13.2 (3.47) | 11.85 (9.45 – 14.35) | 12.41 (4.23) | 0.274 |
| **O<sub>2</sub> Pulse Percent Predicted (%)** | 86 (79.25 – 95) | 89.45 (19.5) | 84 (71 – 101) | 87.3 (25.1) | 0.629 |
| **Heart Rate Reserve (beats per min)** | 18 (12 – 25) | 20.62 (13.31) | 12 (7 – 20) | 12.84 (9.33) | < 0.001* |
| **2-minute Heart Rate Recovery (beats per min)** | 46 (40 – 54) | 45.5 (10.10) | 37.5 (30 – 45) | 37.38(11.74) | < 0.001* |

PEX: Pectus Excavatum, IQR: Interquartile range, \* Statistically Significant

**Table 4.**
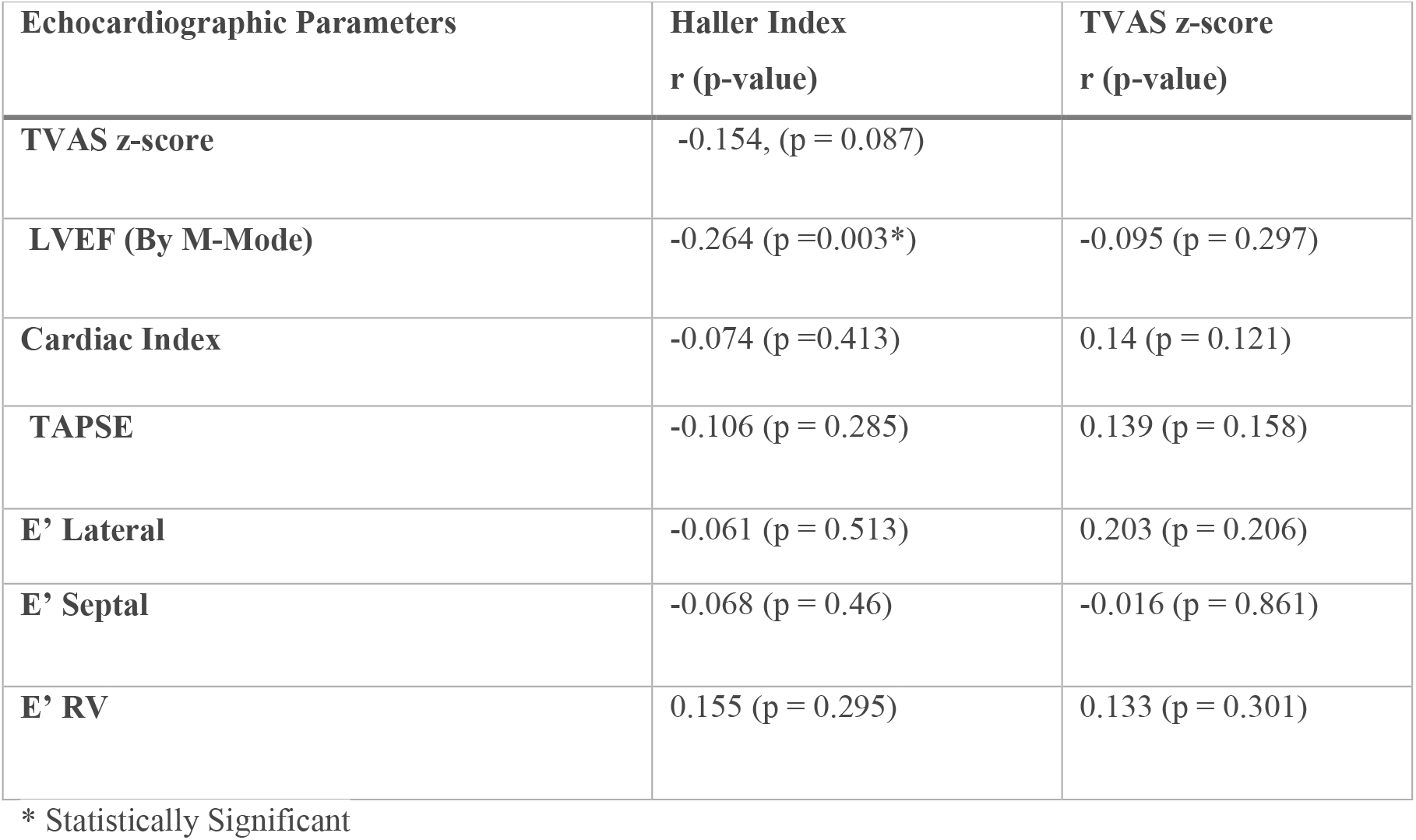
Spearman’s rank correlation between echocardiographic markers and severity of pectus by Haller Index. The TVAS was also specifically evaluated for any correlation with other echocardiographic parameters.

The measured resting LVEF by M-mode was similar and within normal limits for both groups [Mild/moderate PEX LVEF 64% (IQR 61-68); Severe PEX LVEF 63% (IQR 58-66)]. There was however a significant negative correlation between LVEF and the severity of PEX (r = -0.264, p = 0.003) (Table 4).

While the resting cardiac index in the mild/moderate PEX group [2.44 ml/m^2^, IQR (1.82 to 2.93)] was higher than the severe PEX group [2.19 ml/m^2^, IQR (1.84 to 2.53)], this was not statistically significant. There was no significant difference in the Tissue Doppler indices or the TAPSE measurements between the groups (Table 3). The RV FAC could not be reliably obtained in the majority of the patients (94 patients, 75%) due to poor acoustic windows caused by the deformity of the anterior chest wall. Hence, it was excluded from evaluation as an accurate RV FAC requires complete visualization of the free wall, apex, and septal walls of the right ventricle in the apical four-chamber view.

### Cardiopulmonary Parameters

The peak V_O2_ and percent predicted peak V_O2_ were normal for age and not significantly different between the two groups (Table 3). However, there were significant positive correlations between the TVAS z-score with peak V_O2_ (r = 0.023, p = 0.01) and TVAS z score with peak V_O2_ percent predicted (r = 0.19, p = 0.04). Though the oxygen pulse at anaerobic threshold was lower in the severe PEX group this was found to not be statistically significant (p = 0.274). There was however, a strong, significant positive correlation between the TVAS z-score and O_2_ pulse (r = 0.2, p = 0.025) i.e. less compression of the tricuspid valve correlated with a better oxygen pulse. The V_E_/V_CO2_ was similar between the groups with no significant difference but yet again, the TVAS z-score had a significant negative correlation with V_E_/V_CO2_ (r = -0.229, p = 0.011). Finally, it was observed that the 2-minute heart rate recovery was abnormal in PEX and worsened with severity [(mild/moderate PEX: 46 bpm (SD 10) versus 37 bpm (SD 12) in severe PEX (p < 0.001)]. The median heart rate reserve was normal [18 bpm (IQR 12-25)] in mild/moderate PEX but worsened in severe PEX [12 bpm (IQR 7-20)]. This was statistically significant (p < 0.001). The severity of PEX was negatively correlated with heart rate reserve and 2-minute heart rate recovery (r = -0.24, p = 0.006, r = -0.44, p < 0.001) as shown in Table 5.

**Table 5.**
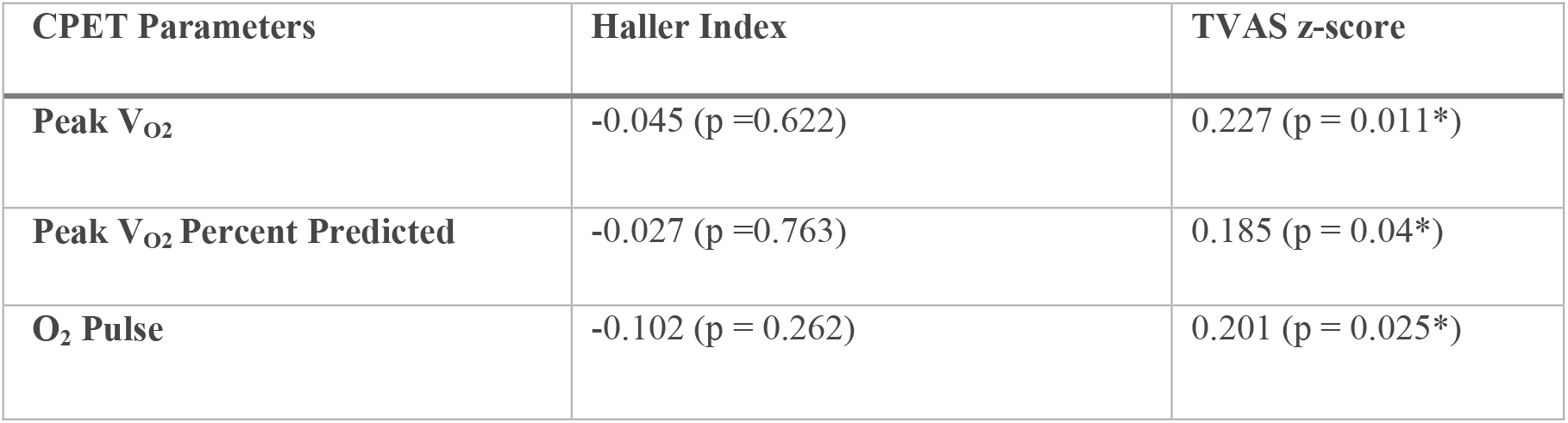
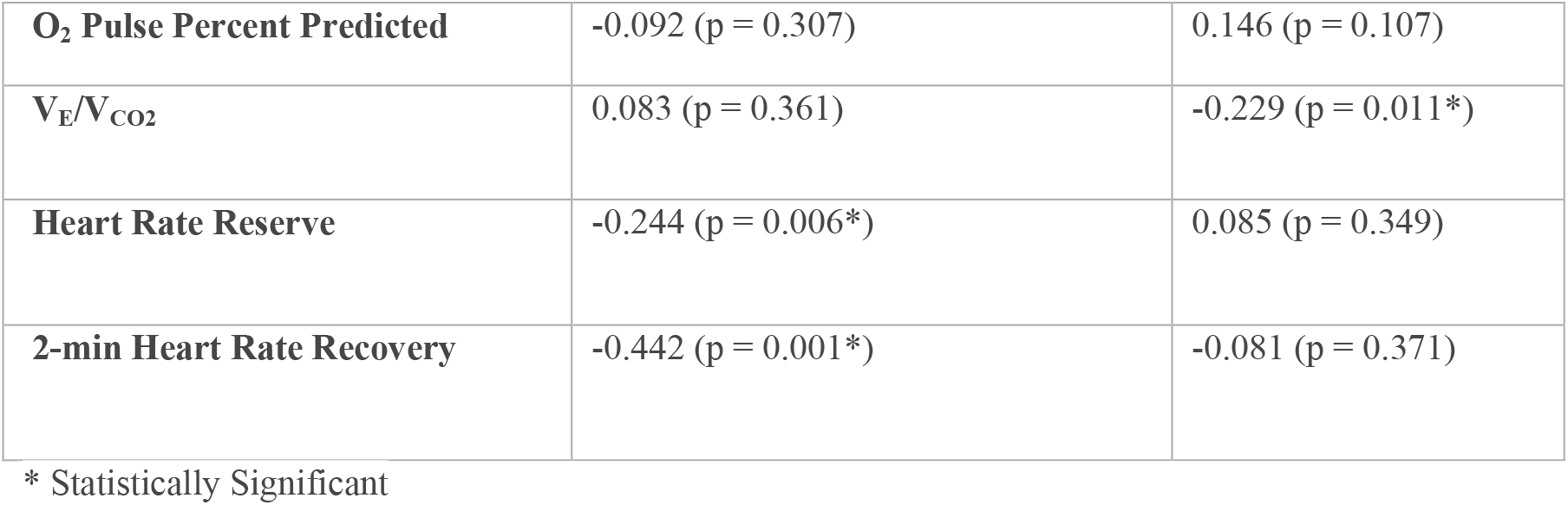
Spearman’s rank correlation comparing severity of pectus by Haller index and TVAS z-score to cardiopulmonary exercise test parameters.

## Discussion

This is the first description of TVAS as a TTE metric for assessing the severity of cardiac compression from pediatric PEX. In this study, the TVAS z-score was found to be lower in those with PEX in comparison to the Detroit normal data set and was significantly lower in the severe PEX group when compared with mild/moderate PEX. That is to say, the tricuspid valve annulus size was smaller when there was significant compression by the PEX (i.e. more severe PEX). Though negatively correlated with the severity of PEX measured by HI, this was not statistically significant. This likely reflects that the degree of cardiac compression is not exclusively related to the severity of the skeletal deformity as represented by HI, but the location of the deformity of the anterior chest wall with relation to the heart is of more importance. A deformity that is lateral to the heart will not impinge on these structures to the same degree as a deformity that is located directly over the heart. This aligns with the findings of a previous study that illustrates that PEX severity by HI correlates with pulmonary limitation but not cardiac compromise (8).This also explains the anecdotal observation from most clinicians that the aerobic capacity is usually normal in PEX.

Our study confirmed that the aerobic capacity as reflected in the peak V_O2_ was not significantly different based on the severity of PEX by HI. However, the lesser the degree of right ventricular compression (as measured by TVAS z-score), the better the aerobic capacity, and this correlation was statistically significant. Similar findings were seen with oxygen pulse which is a marker of stroke volume with exercise (at anaerobic threshold). It correlated with TVAS z-score but not with the severity of PEX by HI. In other words, as the degree of right ventricular compression from PEX increases (not necessarily HI), the tricuspid valve annulus size decreases and the cardiac output during exercise decreases. Other studies have shown a decrease in aerobic capacity and cardiac output with severe HI but have not identified the location of PEX as a factor causing right ventricular compression (19,25).

There was also a significant negative correlation between the ventilator efficiency (as measured by V_E_/V_CO2_) and the TVAS z-score. This indicates that as the tricuspid valve annulus decreases in size from right ventricular compression, the V_E_/V_CO2_ increases (reflecting decreased ventilator efficiency), which may reveal the physiologic interactions of the deformation of the anterior chest wall and restriction of the functional cardiopulmonary system. All of these findings highlight the importance of location of PEX in relation to the heart (echoed by TVAS z-score) over just the skeletal severity of PEX as measured by HI. This may explain why some patients experience symptoms out of proportion to the degree of PEX.

Of note, certain TTE markers were not able to be reliably obtained due to limited acoustic windows caused by the PEX deformity. For example, 74% of the available echocardiograms did not have reliable RV FAC due to incomplete visualization of the free wall, apex, and septal walls of the right ventricle in the apical four-chamber view. Though there was a significant negative correlation between cardiac function as measured by LVEF and the severity of PEX by HI, this was of no clinical significance as it was within the normal range in both groups. In addition, there was no significant correlation between severity of PEX and other TTE indices such as Tissue Doppler indices and TAPSE. This also explains why most TTEs for PEX are reported as “normal” by cardiologists. In a condition like PEX where the acoustic windows are suboptimal due to the extrinsic compression of the right ventricle by the PEX, TVAS proved to be a reproducible metric between cardiologists as reflected in the inter-observer variability. There are adult studies evaluating strain and other novel metrics by TTE and trans-esophageal echocardiography (10,26,27). Advanced imaging such as cardiac magnetic resonance imaging (cMRI) can provide better assessment of these right sided structures (25, 28). However, due to cost and availability, cMRI is not routinely performed in PEX evaluation at most centers. The fact that TVAS measurement is a part of a routine TTE evaluation as opposed to a novel modality that may not be reproducible among general cardiologists, it appears to provide a pragmatic tool and marker for cardiopulmonary distortion especially in a patient with borderline skeletal severity of PEX where they may not qualify for surgery but are symptomatic.

There are no published guidelines on the TTE interpretation of pediatric or adult patients with PEX. Several attempts have been made to characterize the echocardiographic findings in PEX (21-23). This study reiterates the importance of ancillary testing in PEX with CPET and TTE and echoes the importance of focused evaluation of specific indices during interpretation of these modalities. The sole use of HI as a metric for severity of PEX does not take into account the location of PEX and the compression of the right ventricle. In fact, a correction factor has been suggested in abnormal chest morphologies as an adjunct to HI (24). Based on the present study, we propose a TVAS z-score < -2 as a guide for right ventricular compression and cardiopulmonary compromise. This should be used as a metric for clinical indications for repair especially when there are symptoms and the HI does not meet the criteria for correction. The use of TVAS by TTE is a useful armament to the clinician in decision making for PEX.

### Limitations

This study is limited by the retrospective design. There was no longitudinal follow up for the patients to compare potential pre- and post-repair changes in TTE and CPET markers. Despite the large number of patients in the study, the other limitation was the lack of cMRI data to compare with the TTE indices as it was cost-prohibitive and not part of the routine institutional protocol for evaluation of PEX.

Future studies should be performed with longitudinal follow up to compare pre- and post-surgical changes for the TVAS z-score and markers of cardiopulmonary function on CPET. In addition, studies with advanced imaging modalities, such as cMRI, should be used to correlate TVAS measurements on TTE, HI, and metabolic markers on CPET.

## Data Availability

This is to certify full availability of all data referred to in the manuscript.

## Conclusions

Due to the physiologic adaptation with time, peak V_O2_ is not affected significantly in PEX. Tricuspid valve annulus size is a good TTE indicator of cardiopulmonary compromise from PEX. With the relative ease of measurement in a standard TTE, the TVAS z-score offers the clinician a reproducible, pragmatic clinical metric for functional cardiac assessment in PEX. This can potentially add to the armament of indices for decision making for PEX repair.

